# Multi-scalar data integration links glomerular angiopoietin-tie signaling pathway activation with progression of diabetic kidney disease

**DOI:** 10.1101/2021.12.21.21267377

**Authors:** Jiahao Liu, Viji Nair, Yi-yang Zhao, Dong-yuan Chang, Felix Eichinger, Emily C. Tanner, Damian Fermin, Keith A. Bellovich, Susan Steigerwalt, Zeenat Bhat, Jennifer J. Hawkins, Lalita Subramanian, Sylvia E. Rosas, John R. Sedor, Miguel A. Vasquez, Sushrut S. Waikar, Markus Bitzer, Subramaniam Pennathur, Frank Brosius, Min Chen, Matthias Kretzler, Wenjun Ju, for the Kidney Precision Medicine Project and Michigan Translational Core C-PROBE Investigator Group

## Abstract

Diabetes is the leading cause of chronic kidney disease. Prognostic biomarkers reflective of underlying molecular mechanisms are critically needed for effective management of diabetic kidney disease (DKD). In the Clinical Phenotyping and Resource Biobank study, an unbiased, machine learning approach identified a three-marker panel from plasma proteomics which, when added to standard clinical parameters, improved the prediction of outcome of end-stage kidney disease (ESKD) or 40% decline in baseline glomerular filtration rate (GFR) in a discovery DKD group (N=58) and was validated in an independent group (N=68) who also had kidney transcriptomic profiles available. Of the three markers, plasma angiopoietin 2 (ANGPT2) remained significantly associated with composite outcome in 210 Chinese Cohort Study of Chronic Kidney Disease participants with DKD. The glomerular transcriptional Angiopoietin/Tie (ANG-TIE) activation scores, derived from the expression of 154 literature-curated ANG-TIE signaling mediators, positively correlated with plasma ANGPT2 levels and outcome, explained by substantially higher TEK receptor expression in glomeruli and higher ANG-TIE activation scores in endothelial cells in DKD by single cell RNA sequencing. Our work suggests that activation of glomerular ANG-TIE signaling in the kidneys underlies the association of plasma ANGPT2 with disease progression, thereby providing potential targets to prevent DKD progression.

## Introduction

Diabetes is a primary cause of chronic kidney disease (CKD) (1). Early identification of patients at high risk of disease progression is critical for effective management to prevent or delay CKD (2). A recent study suggests that biomarker testing when combined with more effective treatments present a cost-effective way to improve patient outcomes in the diabetic population (3). However, routinely used clinical measures such as estimated glomerular filtration rate (eGFR) and albuminuria have limited sensitivity and specificity for disease prognosis (4; 5). Advances in kidney research over the past decade have revealed multiple biomarker candidates based on transcriptomic (6; 7), proteomic (4) and metabolomic features (8-13) associated with progression of DKD. Due to ease of sample processing, measurement and quantification methods, non-invasive protein markers showing significant association with disease progression are attractive for clinical applications (14-18). However, statistically significant association between biomarker levels and the outcome of interest is only a first step.

A clear underpinning of molecular mechanisms connecting the biomarker with clinical outcome is particularly important when biomarkers are not directly measured in the disease-affected tissue or organ, but rather in body fluids, such as serum or plasma. The unclear origin of these circulating markers raises concerns about the relevance of these markers to kidney pathophysiology. For example, 17 proteins enriched for TNF receptor superfamily members showed strong association with risk of ESKD in patients with DKD (14), where TNFR 1 and 2, despite robust association with ESKD, were likely not produced by the kidney (19). A better understanding of the molecular mechanisms underlying the association of the biomarker with long-term clinical outcomes is needed to prioritize biomarkers for further validation, support potential clinical application for risk stratification and reveal putative therapeutic targets.

Kidney biopsy transcriptomic profiling can help bridge the gap between circulating factors and kidney function impairment. Histopathological and molecular analysis of human kidney biopsy tissue has been the cornerstone of kidney disease diagnosis and prognosis and has provided insights into disease pathophysiology (20-22). Genome-wide transcriptomic profiling of kidney biopsy samples from humans and animal models have helped define disease-associated pathways and better understand DKD pathogenesis (23-25). Recent development of single cell RNA sequencing (scRNAseq) allows comprehensive transcriptomic profiling of kidney biopsies from healthy and disease at single cell-level resolution, providing valuable insights into molecular pathways previously underappreciated due to cellular heterogeneity (26; 27).

In this study, by integrating plasma proteomics and kidney bulk- and single cell-transcriptomic data, we aimed to identify specific circulating markers and delineate the intrarenal signaling cascade that may mediate the association between the circulating biomarker and DKD progression.

## Methods

The overall workflow of the study is depicted in Figure 1.

**Figure 1.**
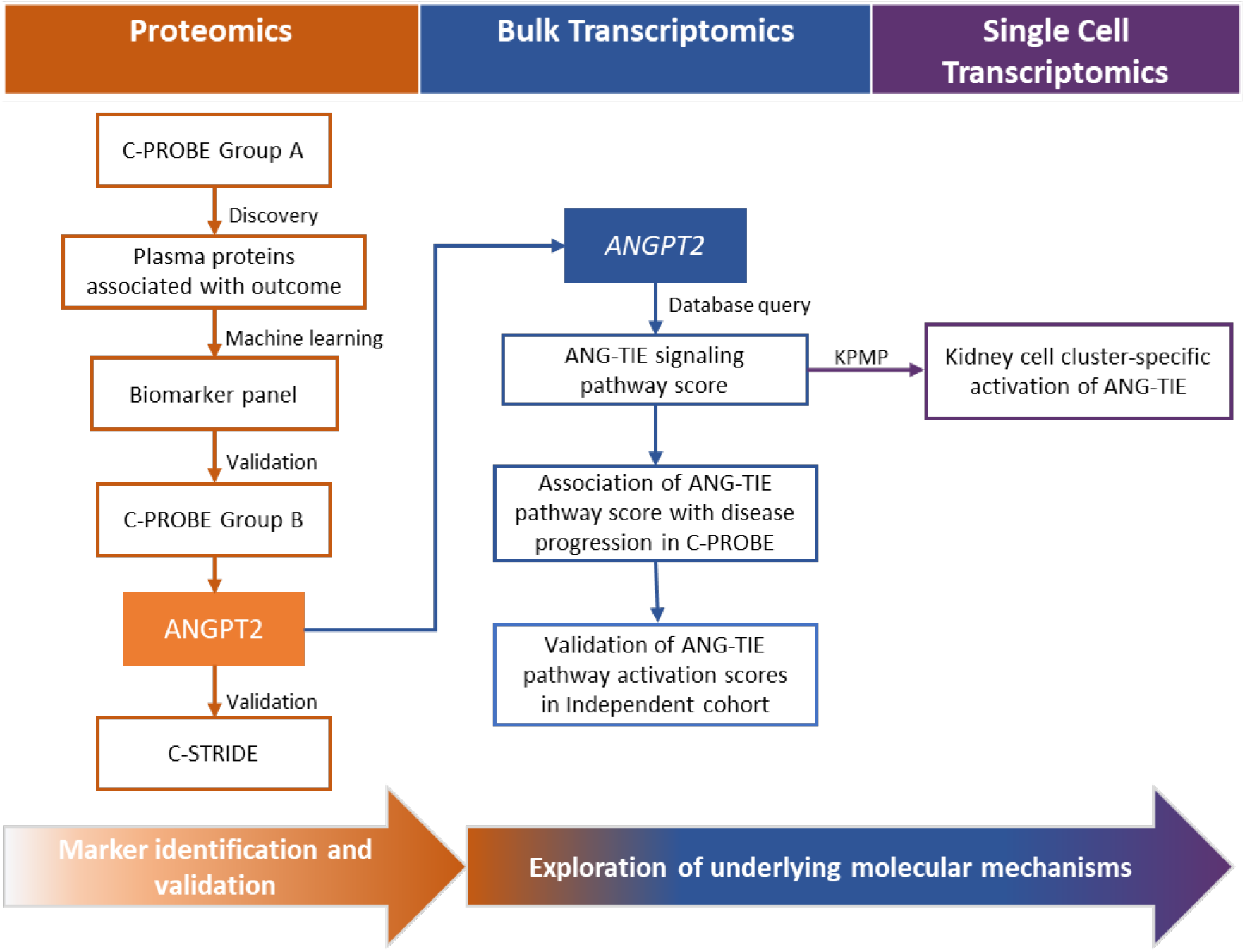
Overall study workflow. SOMAScan plasma proteomics data from C-PROBE group A was used to identify protein markers associated with DKD progression. A three-marker panel that jointly predicted DKD progression was constructed using a machine learning method and validated in plasma proteomics data from C-PROBE group B. Of the three markers, ANGPT2 was further validated using ELISA in the C-STRIDE cohort. To understand the potential biological mechanisms underlying the association of ANGPT2 with kidney outcomes, an ANG-TIE signaling cascade was curated and quantified using bulk transcriptomic data. The associations of kidney ANG-TIE pathway activation scores with circulating ANGPT2 levels and kidney outcomes were evaluated, and the latter validated in an external cohort of patients with DKD (GSE142025). Cell typespecific pathway activation was determined in key kidney cell types using single cell RNAseq.

### Study population

The Clinical Phenotyping and Resource Biobank Core (C-PROBE) is a multiethnic, prospective observational cohort of patients with CKD stages I-IV, collecting clinical information and biospecimens at enrollment and yearly thereafter at six sites in the United States (http://kidneycenter.med.umich.edu/cprobe)(28). The C-PROBE discovery group (A) included patients with CKD and diabetes who had three or more eGFR measurements during the follow-up period and had plasma samples available from time of enrollment. Patients with baseline eGFR < 30 mL/min/1.73m ^2^ were excluded based on advanced disease. eGFR was estimated using the CKD-EPI equation (29). Albuminuria was assessed by Albuwell Hu ELISA Kit (Ethos Biosciences, Newtown Square, PA). eGFR slopes were calculated using a linear mixed-effects models (28). Patients with eGFR slopes less than -3%/year and above 3%/year were matched for major confounders including age, sex, race, and baseline eGFR. The independent C-PROBE validation group (B) included participants with kidney biopsy transcriptomic profiles.

The Chinese Cohort Study of Chronic Kidney Disease (C-STRIDE) is a multicenter prospective cohort of CKD patients (30). Participants with: (i) diagnosis of diabetes mellitus or had fasting blood glucose ≥7 mmol/L; (ii) eGFR<60 and >30 mL/min/1.73m^2^ or uACR >30mg/g for at least three months; (iii) had two or more available eGFR measurements during follow-up and (iv) had plasma samples available from the baseline visit were included. eGFR was estimated using the Modification of Diet in Renal Disease equation (31).

### Kidney outcomes and covariates

The outcome was defined as ESKD or more than 40% reduction from baseline eGFR, whichever occurred first. Participants who reached the outcome were defined as progressors. Covariates included age, gender, race, eGFR and uACR documented at study enrollment. Participants missing key covariates were not included in the analyses.

### SOMAScan Measurement

SOMAmer-based proteomic assay using the SOMAscan 1.3K Plasma kits (19; 32) per standard experimental and data analysis protocols (33) were performed by the Genome Technology Access Center at the McDonnell Genome Institute at Washington University School of Medicine. Detailed protocol and analysis are included in supplementary methods.

### Measurement of ANGPT2 using ELISA

Human Angiopoietin-2 Quantikine ELISA (R&D Systems, DANG20) was used according to the manufacturer’s instructions. Plasma samples (1:10) and standards were assayed in duplicate. Three quality controls with high, medium, and low concentrations were included in each plate to control for inter-plate variability. Absorbance at 450 nm was read with the VersaMax Microplate Reader (Molecular Devices) and results were calculated with the SoftMax Pro software. The quality control measures of % recovery mean and % coefficient of variation (CV) were within acceptable ranges and the inter-assay coefficient of variation was below the 4.0% threshold.

### Statistical analysis

The descriptive statistics are presented as median (interquartile range) or proportions. Cox proportional hazard model was used to determine the association of biomarker with composite endpoint. Statistically significant biomarkers (P<0.05) in a univariate model were used as input for the machine learning approach. Lasso Cox method (34) was used for feature reduction (details in supplementary methods). A multivariate Cox model based on the markers selected by lasso was constructed to evaluate the performance of each marker. Model performance was further evaluated using C statistic (35) and time dependent AUC (area under the curve) for survival data (36). The performance of two competing models were compared using delta C statistic based on a perturbation-resampling method (35), and the Likelihood Ratio (LR) test for its sensitivity in capturing the differences in prediction ability. R package glmnet (37; 38) was used to construct the lasso Cox model. Cox proportional model was constructed using R package survival (Therneau, Grambsch, 2020), C statistic was calculated using R package survC1 (35), time dependent AUC was calculated using R package timeROC (36). All analyses were completed using R version 3.6.2. (R Core Team, 2019).

### Gene expression data analysis

Gene expression profiling from micro-dissected kidney biopsies was performed as previously described (39) using Affymetrix 2.1 ST chips (Affymetrix, Santa Clara, CA). Probe sets were annotated to Entrez Gene IDs using custom CDF V.19 generated from the University of Michigan Brain Array group (40). Expression data were normalized across genes using robust multi-array average (RMA) (41). The Cel files can be accessed through the Gene Omnibus, GSE180395 (https://www.ncbi.nlm.nih.gov/geo/query/acc.cgi?acc=GSE180395).

RNAseq data, GSE142025, fastq files were downloaded from GEO (42; 43), reads quality inspected using FastQC, qualitied reads were mapped to human reference genome GRCh38 using STAR 2.5.2b (44). The mapped reads were then examined for distribution across exons, introns, UTR, and intergenic regions using Picard Tools (https://broadinstitute.github.io/picard/). Gene level expression quantification was performed using HTSeq (45), and the resultant count data were normalized in voom (46). PCA and hierarchical clustering were used to identify and remove samples with abnormal expression profiles due to technical issues, and the mapping statistics obtained from STAR.

scRNASeq data from 10 patients with DKD and 18 living kidney transplant donors (LD) were downloaded from Kidney Precision Medicine Project (KPMP) kidney tissue atlas (https://atlas.kpmp.org/repository) and processed according to the KPMP single cell protocol as previously described (18; 47). In the dot plot analysis, the size of the dots represents the percentage of cells expressing *TEK* (*TEK* (+) cells). *TEK* (+) expression was defined based on greater than zero (*TEK* > 0) normalized gene expression, remaining cells were considered *TEK*(-).

*TEK/TIE2* gene expression in heathy donors were extracted from Nephroseq database (https://www.nephroseq.org/) using the Lindenmeyer normal kidney tissue panel (48).

### Gene set curation, network visualization and functional module detection

The union of ANG-TIE signaling pathway genes curated from three databases, namely the “Angiopoietin receptor Tie2-mediated signaling pathway” from the Pathway Interaction Database (PID) (49), the “Tie-2 signaling pathway” from the REACTOME database (50) and “TIE2-angiopoietin signaling pathway” from the NETPATH database (51) provided a set of 154 genes (Figure S1). Cytoscape (52) was used to visualize the genes and their database sources. Community clustering in HumanBase database (53) was applied to the ANG-TIE signatures within the kidney functional network to identify cohesive gene modules.

### Creation of gene-set based pathway activation score

Log2 transformed gene expression at bulk transcriptome level was used to compute Z-scores, and the average of Z-scores of the 154 pathway gene signature was used to generate an ANG-TIE pathway activation score for each participant (54). The AddModuleScore function in R package Seurat (55; 56) was used to generate ANG-TIE pathway scores at the single cell cluster level. The pathway scores were also grouped into glomerular (podocytes, mesangial cells, vascular smooth muscle, and endothelial cells) and tubulointerstitial (proximal tubular, descending and ascending thin limb, think ascending limb, connecting tubular, intercalated and principal) cells based on cellular origin.

## Results

### Circulating biomarkers associated with DKD progression

Fifty-eight C-PROBE participants were included in the discovery group (C-PROBE group A) and their baseline as well as follow-up characteristics are summarized in Table 1. Mean age at enrollment was 49.54 (13.74) years. Participants had impaired baseline kidney function, with mean eGFR of 49.54 (13.74) ml/min/1.73m^2^, and median uACR 167.4 (IQR =683.09) mg/g. After 4.4 (IQR=3) years of follow-up, 28 of 58 patients reached the outcome.

**Table 1.** Basic demographics and clinical characteristics of participating patients from C-PROBE and C-STRIDE.

|  | C-PROBE<br>Discovery group A<br>(n=58) | C-PROBE<br>Validation group B<br>(n=68) | C-STRIDE<br>(n=210) |
| --- | --- | --- | --- |
| **At baseline** |  |  |  |
| Age (years) | 59.90 (11.70) | 43.57 (15.35) | 58.52 (10.61) |
| Female (%) | 33 (56.9) | 41 (60.3) | 80 (47.3) |
| Race black (%) | 28 (48.3) | 13 (19.1) | NA |
| eGFR (ml/min per 1.73m<sup>2</sup>) | 49.54 (13.74) | 65.61 (34.43) | 57.22 (30.62) |
| uACR (mg/g) | 167.4 [683.09] | 816.2 [1680.8] | 595.0 [1364.9] |
| **During follow-up** |  |  |  |
| Length of follow-up(years) | 4.4 (3.0) | 3.6 (2.8) | 1.9 (2.3) |
| Numbers reaching outcome (%) | 28 (48.3) | 29 (49.2) | 74 (35.2) |

Data are shown as mean (SD) or median [interquartile range, IQR] for continuous variable or n (proportions) for categorical variable.
Outcome was defined as time to composite event of developing end stage kidney disease (ESKD) or loss >40% of the baseline eGFR, whichever occurs first.
Data were complete except uACR missing for 1, 4 and 7 individuals in C-PROBE group A, C-PROBE group B, and C-STRIDE, respectively. Time of follow-up is missing for 5 people in C-PROBE group B.
Abbreviations: eGFR- estimated glomerular filtration rate; uACR- urinary albumin-to-creatinine ratio; NA – not applicable.

Using the Cox proportional hazard model, 84 plasma proteins of 1301 measured on the SOMAScan platform were univariately associated with time to reaching the composite endpoint (p<0.05, Figure 2a, Supplementary Table S1). Epidermal growth factor receptor (EGFR), apolipoprotein A1 (APOA1), cathepsin V (CTSV), c-type lectin domain family 4 member M (CLEC4M) and TNF receptor superfamily member 1A (TNFRSF1A) were among the markers that showed the strongest associations with outcome. Many of the 84 proteins were intercorrelated, with Pearson correlation coefficients ranging from -0.63 to 0.96 (Figure S2a). Correlation matrix of those proteins ordered by hierarchical clustering showed distinct correlation patterns (Supplementary Figure S2b) suggesting that those proteins may represent different aspects of pathogenesis of DKD progression. The intercorrelated expression pattern suggested the need for a feature selection approach to reduce excessive multicollinearity.

**Figure 2.**
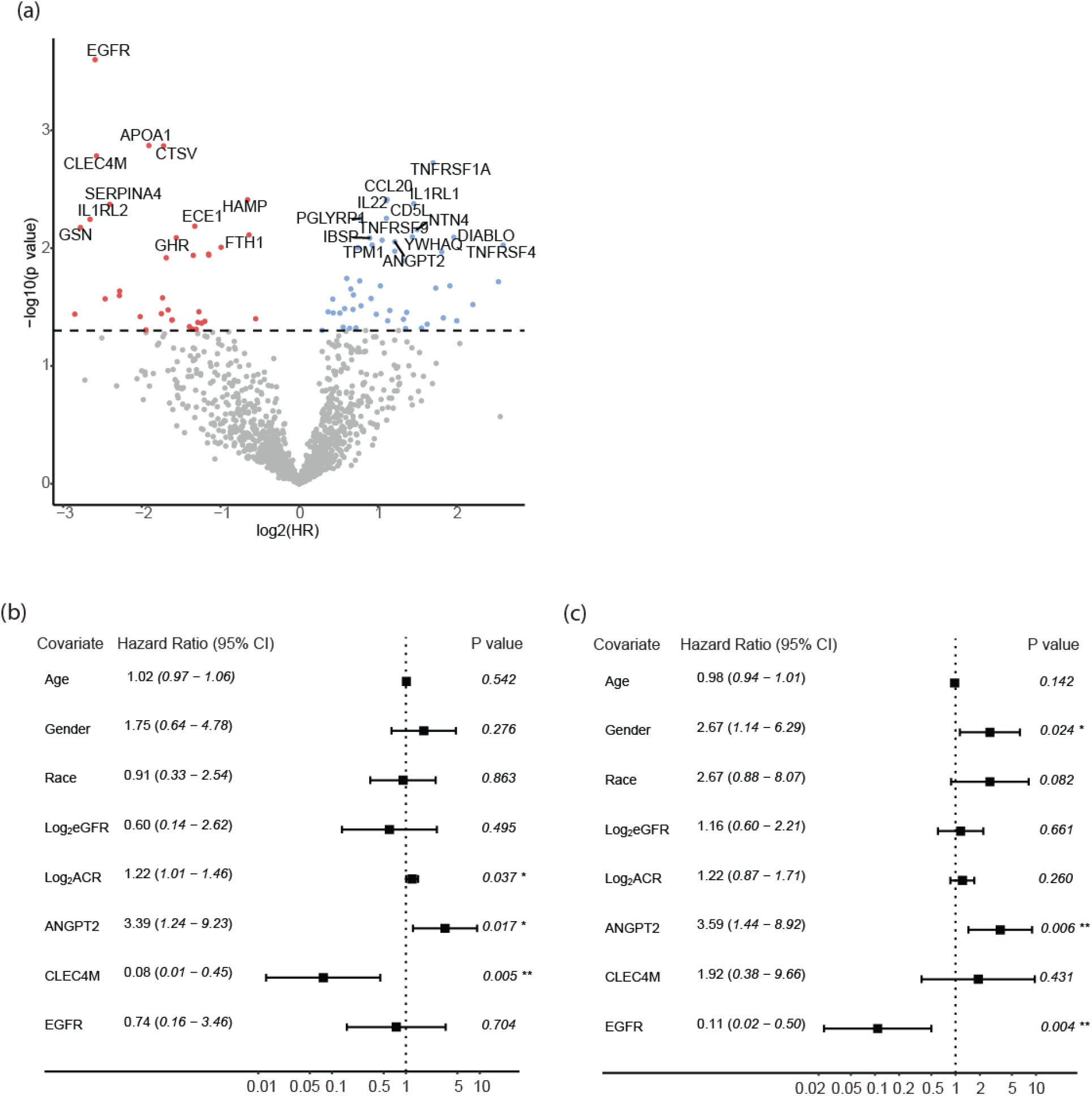
Identification and validation of the biomarker panel for DKD progression. (a) Volcano plot demonstrating the associations of circulating proteins with risk of progression to the composite outcome. Hazard ratio are in log_2_ scale(x-axis) and statistical significance of associations are in log_10_ scale (y axis) are presented. Forest plot of the multivariate Cox proportional hazard model in C-PROBE discovery Group A (N=58) (b) and validation group B (N=68) (c). *-p<0.05, **-p<0.01

### Predictive biomarker panel for DKD progression

To reduce dimensionality and construct an actionable marker panel that could jointly predict DKD progression, a feature selection procedure (57-59) was applied to the 84 proteins (cross-validation curve provided in Supplementary Figure S2c). A panel of 3 biomarkers (EGFR, CLEC4M and ANGPT2) was selected using this procedure. The markers were evaluated together with clinical variables (Figure 2b), with CLEC4M (Hazard ratio (HR)=0.076, p=0.005) and ANGPT2 (HR=3.39, p=0.02) remaining statistically significant in the multivariate Cox proportional hazard model. Addition of the biomarker panel significantly improved the prediction of DKD progression (LR test p =0.003) over clinical variables alone (model 0), with C statistic improving from 0.728 to 0.791 for the joint model including clinical variables and the selected biomarker panel (model 2, Table 2). Time-dependent ROC (receiver operating characteristic) curve for the Cox model (truncated at 5 years) also demonstrated that the lasso selected biomarker panel could improve the prediction when added to clinical variables, with an increase in AUC from 0.704 to 0.806 (Figure S3).

**Table 2.** Incremental value of the biomarker panel in prediction of composite endpoint of ESRD or 40% reduction in eGFR.

|  | C statistic<br>(CI) | delta C statistic<br>(CI)<sup>a</sup> | P value<br>(LR test)<sup>a</sup> |
| --- | --- | --- | --- |
| **Group A (discovery)** |  |  |  |
| Model 0 | 0.728 (0.583-0.873) |  |  |
| Model 1 | 0.773 (0.671- 0.874) |  |  |
| Model 2 | 0.791 (0.662,0.920) | 0.063 (-0.031-0.157) | 0.003 |
| **Group B (validation)** |  |  |  |
| Model 0 | 0.671 (0.531-0.811) |  |  |
| Model 1 | 0.703 (0.596 -0.810) |  |  |
| Model 2 | 0.787 (0.666-0.907) | 0.116 (-0.002-0.234) | 0.0004 |

Model 0 covariates: Age, gender, race, eGFR and uACR
Model 1 covariates: three biomarkers EGFR, CLEC4M and ANGPT2
Model 2 covariates: Age, gender, race, eGFR, uACR and three biomarkers EGFR, CLEC4M and ANGPT2.
<sup>a</sup>delta C statistic and P value are the results comparing model 2 and model 0.
Abbreviations: CI-confidence interval; LR test- likelihood ratio test.

### Validation of the additive value of the biomarker panel in an independent group of patients

To evaluate the performance of the 3-marker panel in an independent group of patients, concentrations of the identified biomarkers were measured by SOMAScan assay in plasma samples of the C-PROBE validation group B (N=68, Table 1). Group B included patients with transcriptomic data derived from kidney biopsies. In comparison with patients from Group A, patients from Group B were younger, had higher eGFR and uACR at baseline, and fewer patients were black. Mean eGFR at enrollment was 65.61 (34.43) ml/min per 1.73m2, and median uACR was 816.2 (1680.8) mg/g. Twenty-nine participants reached the outcome after a median follow-up of 3.6 years.

The three-marker panel showed significant improvement in prediction of progression to composite endpoint with an increase of C statistic from 0.671 (model 0, clinical model) to 0.787 for model 2 (the joint clinical and biomarker model) with a significant LR test p value of 0.0004 (Table 2) in this validation group. Of the three markers, ANGPT2 remained statistically significant with HR of 3.59, namely one unit increase of ANGPT2 (in log_2_ scale) was significantly associated with 259% increased risk of progression given the other covariates withheld (p=0.006, Figure 2c).

### Validation of ANGPT2 concentration using enzyme-linked immunosorbent assay (ELISA)

ANGPT2 stood out as a significant predictor in both the discovery (C-PROBE group A) and validation group (C-PROBE group B) after adjusting for age, sex, race, eGFR and uACR, and the other two biomarkers (Figures 2b and c). To confirm concentrations measured by SOMAScan with an independent technology, ANGPT2 in plasma samples from C-PROBE group A were measured using ELISA. The ANGPT2 concentrations measured using these two technologies were strongly correlated, with a correlation coefficient of 0.91 (p=2.58e-23, Figure 3a). The association of ANGPT2 with outcome was replicated (HR: 2.11 (1.30-3.41)). When stratified by ANGPT2 quartiles measured by ELISA, the risk of reaching the outcome increased by 8.04-fold (95% CI: 2.13-30.38, p=0.002) in patients from quartile 4 compared to quartile 1 (Table S2).

**Figure 3.**
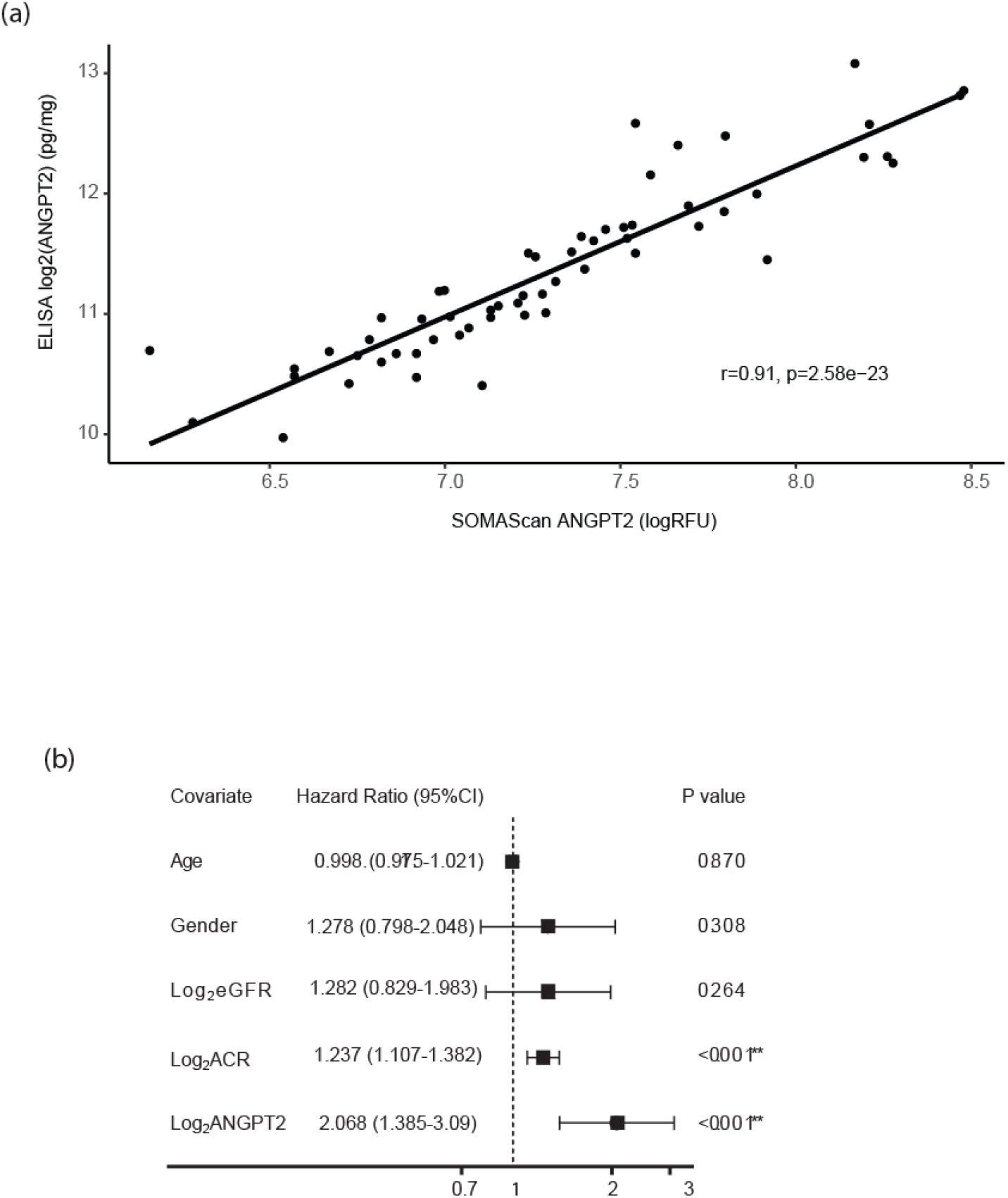
Validation of the significant association of plasma ANGPT2 with the risk of composite endpoint in C-STRIDE cohort. (a) ANGPT2 concentrations measured by two different assay platforms, SOMAscan and ELISA, are significantly correlated with each other in patients from C-PROBE cohort A; (b) Forest plot showing significant association of ANGPT2 with composite endpoint in multivariate Cox proportional hazard model in DKD patients from C-STRIDE cohort where ANGPT2 concentrations were measured using ELISA. *-p<0.05, **-p<0.01

### Validation of the association of ANGPT2 with composite endpoint in an external cohort

The association of ANGPT2 with composite outcome was further validated in an independent, external, multi-center cohort of patients with DKD, the Chinese Cohort Study of Chronic Kidney Disease (C-STRIDE). The baseline characteristics of the C-STRIDE patients (N=210, Table 1), including the means of age and eGFR, and median of uACR were similar to C-PROBE group A and B. Length of follow-up was shorter in patients from C-STRIDE. Higher baseline concentrations of ANGPT2, measured using ELISA, were associated with increased risk of the composite endpoint, the estimated HR (95% CI) was 2.09 (1.40 to 3.12), adjusted for age, gender, eGFR, and ACR (Figure 3b). Thus, with one unit increase of ANGPT2 (in log_2_ scale) and the other covariates withheld, the hazard of disease progression would increase by 109%, in C-STRIDE.

### ANGPT2-associated gene network in the kidney

The mechanistic role of circulating ANGPT2 on kidney disease progression were investigated by integrating transcriptomic data of C-PROBE group B participants. To understand the ANGPT2 downstream signaling cascade in the kidney and its association with outcome, we generated the 154 ANG-TIE signaling network gene list (Table S3, Figure 4a). Community clustering of the 154 genes through Humanbase kidney functional network analysis (53) resulted in three gene cluster modules (Figure S4) that contained key processes closely related to transmembrane receptor protein tyrosine kinase signaling (in both M1 and M2), regulation of cellular response to insulin stimulus and ephrin receptor signaling pathway (M1), superoxide metabolic process and positive regulation of cell motility (M2), and cellular response to oxidative stress and regulation of apoptotic process (M3).

**Figure 4.**
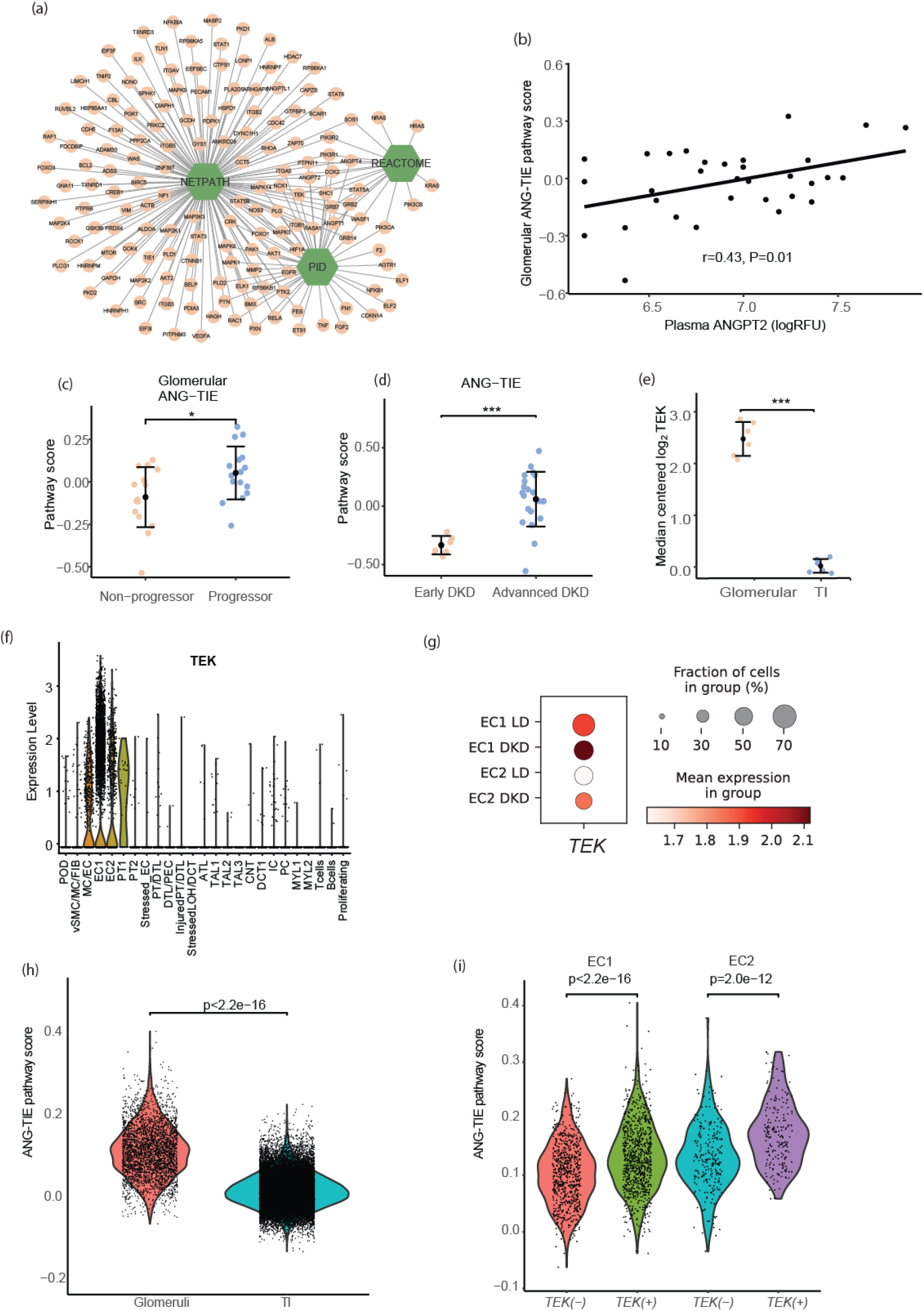
ANG-TIE signaling pathway in micro-dissected transcriptome and KPMP scRNAseq data. (a) Network visualization of the literature curated ANG-TIE signaling cascade network. (b) Association of plasma ANGPT2 with glomerular ANG-TIE signaling pathway score in (n=32). (c) The glomerular ANG-TIE signaling pathway score is significantly higher in progressors compared to non-progressors in C-PROBE cohort B (n=32). (d) The ANG-TIE signaling pathway score is significantly higher in patients with advanced DKD compared to early DKD from GSE142025 dataset (n=28). Definition of early and advanced DKD is adapted from the original paper and outlined in the methods section. (e) *TEK* gene expression is significantly higher in glomeruli than in tubulointerstitia (Data derived from Lindenmeyer normal kidney tissue panel in Nephroseq). (f) ScRNAseq analysis of a combined analysis of 56906 cells from KPMP datasets demonstrate *TEK* gene expression is enriched in endothelial cells (EC), followed by MC/EC cluster, sporadic expression was also observed in PT1 cells. (g) Dot plot of *TEK* expression (higher expression indicated by increase in color intensity of the dots) in patients with DKD compared to living donors. (h) Violin plots showing ANG-TIE pathway activation scores are significantly higher in glomerular cells compared to tubulointerstitial cells of kidney biopsy samples from patients with DKD. (i) Violin plots showing the ANG-TIE pathway activation scores are significantly higher in cells expressing higher *TEK* (*TEK*-H) cells compared to cells expressing low level of *TEK* (*TEK*-L), in both endothelial cell clusters of DKD patients. For figure c, d and e, student t test was used and for panels h and i, Mann–Whitney U test was used to compare the difference between groups.

### Association of ANG-TIE activation score with kidney outcome

The ANG-TIE pathway activation scores were calculated for every C-PROBE group B participant by aggregating the expression levels of the 154 pathway genes, in the glomerular and tubulointerstitial compartments data. The association of ANG-TIE pathway activation scores with plasma ANGPT2 levels were evaluated in a compartment-specific manner. For patients with both glomerular transcriptomic data and plasma SOMAscan data (N=32), significant positive correlation between plasma ANGPT2 levels and glomerular ANG-TIE activation scores (r=0.43, p=0.01, Figure 4b) were observed. No significant correlations were observed between tubulointerstitial ANG-TIE pathway activation scores and plasma ANGPT2 levels (N=25, r=0.16, p=0.82, Figure S5a).

The glomerular ANG-TIE pathway activation score was significantly higher in progressors (those who reached composite endpoint during follow-up) compared to non-progressors (the rest of C-PROBE group B) (p=0.02, Figure 4c); no significant differences were observed in tubulointerstitial ANG-TIE scores between these two groups (p=0.51, Figure S5b).

To test the generalizability of these findings, we downloaded the kidney cortex transcriptomic data from 28 patients with biopsy-proven diabetic nephropathy (DN) (43). Twenty-seven samples passed the quality control criteria and were divided into two groups (43), early (n=6) and advanced DN (n=21). Early DN was defined as uACR between 30 and 300 mg/g, eGFR <90 mL/min/1.73 m^2^, whereas advanced DN was defined as uACR >300 mg/g, eGFR <90 mL/min/1.73 m^2^. Significantly higher ANG-TIE pathway activation scores were observed in the advanced compared to early DN group (p=9.98e-7, Figure 4d) in this independent, external data set.

### Compartment- and cell cluster-specific ANG-TIE signaling pathway activation in the kidney

To uncover the link between glomerular ANG-TIE scores, plasma ANGPT2 and disease outcome, we evaluated the mRNA expression of *TEK*, the receptor of ANGPT2, in micro-dissected glomeruli and tubulointerstitia from biopsies of 6 healthy donors using Nephroseq data from Lindenmeyer normal kidney tissue panel (48). *TEK* expression was significantly higher in glomeruli compared to tubulointerstitia (Figure 4e).

Further, ScRNAseq data from kidney biopsies of 10 participants diagnosed with DKD were accessed through the KPMP kidney tissue atlas (Table S4, https://atlas.kpmp.org/repository) (47). The previously published scRNAseq data from 18 living donors (LDs) were the controls (18). A combined analysis of 56906 cells from these two data sets demonstrated that the TEK receptor was specifically expressed in endothelial cell clusters (Figure 4f, Figure S6a), and the average intensity of *TEK* is significantly higher in patients with DKD compared to living donors (Figure 4g, figure S6b and S6c).

ANG-TIE pathway scores were calculated at the single cell level. Comparison of the pathway scores grouped by cells from glomerular and tubulointerstitial compartments (glomeruli vs tubulointerstitia) indicated a more active ANG-TIE signaling in glomeruli compared to tubulointerstitia (Figure 4h, p<2.2e-16), consistent with a glomerulus-enriched TEK receptor expression pattern (Figures 4e).

About half the cells in the endothelial cell clusters (EC1 and EC2) in the DKD samples expressed *TEK* (*TEK* (+) cells). The level of ANG-TIE activation score was significantly higher in *TEK* (+) compared to *TEK* (-) endothelial cells in DKD (Figure 4i).

## Discussion

Multiple circulating protein biomarkers have been previously reported as strongly associated with progression of DKD (4-18). However, the mechanistic links of these molecules to cellular events in the kidney that would explain their association with kidney damage remain unclear. This study identified and validated a plasma marker panel, including ANGPT2, that is associated with long term clinical outcomes, but more importantly, provides insights into a plausible molecular signaling cascade underlying this association through multi-scalar data integration.

An aptamer-based proteome assay, enabled identification of plasma protein markers that are associated with DKD progression, including previously reported DKD biomarkers, such as β2-microglobulin (B2M) (60), tumor necrosis factor (TNF)-receptor superfamily members TNFRSF1A, TNFRSF1B, TNFRSF9 and TNFRSF4 (19; 61-63), CD27 antigen (CD27) (64; 65)

CCL2/MCP-1 (66), and ANGPT2 (67). Novel candidate biomarkers involved in disease functions, such as apoptosis (diablo IAP-binding mitochondrial protein (DIABLO) (68), HtrA serine peptidase 2 (HTRA2) (69)); inflammation (interleukin 1 beta (IL1B) (70), interleukin 22 (IL22) (71)); and angiogenesis (fibroblast growth factor 2 (FGF2) (72), notch receptor 1 (NOTCH1) (73)) were also identified. Unbiased machine learning methods narrowed the list to a three-biomarker panel (EGFR, CLEC4M, ANGPT2) that improved prediction of the composite endpoint and amongst these, ANGPT2 persisted across three independent DKD groups.

ANGPT2 is a ligand for the endothelial TEK receptor tyrosine kinase (74). It inhibits TEK/TIE2 phosphorylation and induces pro-inflammatory and pro-fibrotic genes (74; 75). ANGPT2 is stored in and rapidly released upon stimulation from endothelial cell Weibel-Palade bodies (76). The release of ANGPT2 into the bloodstream is triggered by hypoxia, inflammation and high glucose levels (77). ANGPT2 is crucial to the induction of inflammation and sensitizes endothelial cells to TNF-α (78). Increased serum or plasma levels of ANGPT2 were initially associated with cardiovascular disorders (74; 79-81) but also found elevated in DN (82). ANGPT2 has been associated with increased risk of kidney function decline in DKD (67) and CKD (83), and mortality in CKD (67; 83; 84). This study, for the first time, revealed that elevated circulating ANGPT2 functions through the TIE2 receptor in intra-renal endothelial cells and thereby activates a downstream pro-inflammatory and pro-fibrotic signaling cascade. We discovered that higher TEK receptor expression in glomeruli is associated with higher glomerular ANG-TIE signaling pathway scores and that the glomerular, not tubulointerstitial, ANG-TIE signaling pathway scores significantly correlated with circulating ANGPT2 levels, arguing for a critical role of glomerular endothelial cell signaling in mediating the association of circulating ANGPT2 with adverse DKD outcomes.

Our findings are consistent with studies showing an essential role for ANG-TIE signaling in the glomerular capillaries in both physiology and disease (85). Although ANGPT2 appears to act primarily as an autocrine antagonistic regulator (76), it could function as a paracrine regulator. For example, podocyte-specific ANGPT2 overexpression in pre-clinical models increased proteinuria and endothelial cell apoptosis (86), caused endothelial glycocalyx breakdown (87) and specifically stimulated endothelial expression of chemokines and adhesion molecules (75).

Potential therapeutics such as Faricimab (88) that manipulate ANGPT2 levels have provided promising results in cancer (89) and retinopathy (90; 91). ANGPT2 or ANG-TIE signaling have yet to be targeted in DKD or CKD. Based on our findings, ANGPT2 or ANG-TIE signaling pathway could also offer therapeutic targets for DKD (92; 93).

The strengths of this study include the replication of the high throughput SOMAscan results with ANGPT2 concentration measured using ELISA, and validation of findings in the independent C-STRIDE cohort. The pathway activation score generated in this study can be used to determine the ANG-TIE pathway activity in kidney biopsies in future clinical studies and preclinical models of kidney disease. Further, this 154 gene set could be a starting point for identifying urinary predictive and pharmacodynamic biomarkers for ANG-TIE associated therapeutics.

Due to the limited sample size of the discovery group A and the large number of proteins measured, the association of circulating proteins with DKD progression was only significant at the nominal p value threshold, with statistical significance lost after multiple testing. This is a well-known limitation of small sample sizes in high-dimensional data analysis (94), that we addressed in several ways. Critically, we confirmed the significant association of ANGPT2 with composite endpoint in an independent group of patients from C-PROBE (group B) and further confirmed the finding in the C-STRIDE cohort, using two different assay platforms. Additionally, since biology is key for a robust biomarker, we validated the association of the ANG-TIE pathway activation score with outcome in an independent, publicly available data set. The verification of the initial discovery in multiple, independent, and external validation groups to reinforce the reliability of the discovery, potentially compensates for the limitations of multiple-testing associated with small sample size, frequently encountered in translational and preclinical research. Nevertheless, further validation using antibody-based immunoassays assays in large and independent study cohorts are warranted to develop ANGPT2 or other candidate biomarkers, identified in this study, as clinical prognostic markers.

In summary, our integrative bioinformatics analysis identified and validated circulating ANGPT2 as a potential prognostic marker for DKD progression and revealed the link between plasma ANGPT2, glomerular ANG-TIE signaling and the DKD progression outcome of ESKD or 40% loss of eGFR. Our findings support further investigations into ANG-TIE signaling targeted therapies, currently being tested in other conditions, to prevent or ameliorate DKD progression.

## Data Availability

Data used in the present study can be made available through appropriate approvals from cohort study committees.
Transcriptomic data were also accessed through GEO: GSE142025,

https://www.ncbi.nlm.nih.gov/geo/query/acc.cgi?acc=GSE180395

https://atlas.kpmp.org/repository

https://www.nephroseq.org/

## Acknowledgements

This work was supported by a grant from the University of Michigan Health System and Peking University Health Sciences Center Joint Institute for Translational and Clinical Research (No. BMU2017JI001). Support provided by the China Scholarship Council (201906370288) during a visit of JL to University of Michigan is acknowledged. This study was also supported, in part, by funding from NIH/NIDDK through the George M. O’Brien Michigan Kidney Translational Core Center, grant 2P30-DK-081943; the Integrated Systems Biology Approach to Diabetic Microvascular Complications grant, R24DK082841; P30DK89503; and JDRF Center for Excellence (5-COE-2019-861-S-B). The Kidney Precision Medicine Project (KPMP), UH3-DK-114907, is a multi-year project funded by the National Institute of Diabetes, Digestive, and Kidney Diseases (NIDDK) with the purpose of understanding and finding new ways to treat chronic kidney disease (CKD) and acute kidney injury (AKI). See Supplemental Acknowledgments for consortium details. C-STRIDE is supported by a grant from China International Medical Foundation-Renal Anemia Fund, the grants from the National Natural Science Found (No. 82070748, 82090020 and 82090021). Dr. Vasquez is supported through funds from the National Institutes of Health, 5UH3DK114870-05. The authors thank Drs. Yuee Wang and Virginia Vega-Warner for their technical support.

## Supplementary Materials

1. Supplementary acknowledgement – KPMP
2. Supplementary methods
3. Supplementary references
4. Supplementary tables:
  - Table S1. Proteins associated with DKD progression.
  - Table S2. Association of plasma ANGPT2 (ELISA) with risk of composite outcome
  - Table S3. ANG-TIE signaling pathway related genes from three database
  - Table S4. Basic clinical data of DKD patients included in the ScRNAseq analysis
5. Supplementary Figures: S1 to S6
  - Figure S1. Curation of an unbiased ANG-TIE signaling gene set.
  - Figure S2. Correlation matrix between the univariate significant plasma proteins and the lasso cross validation curve.
  - Figure S3. Time-dependent ROC curve truncated at 5 years for clinical model and the joint clinical and biomarker model.
  - Figure S4. Functional characterization of the ANG-TIE signaling network in the kidney.
  - Figure S5. Association of tubular ANG-TIE signaling pathway score with plasma ANGPT2 level and kidney outcome (n=25).
  - Figure S6. TEK gene expression in KPMP single cell data: DKD and living kidney donors (LD).

## Abbreviations

TI: tubulointerstitial
ATL: ascending thin loop of Henle
CNT: connecting tubule
DCT: distal convoluted tubule
DTL: descending loop of Henle
EC: endothelial cell
FIB: fibroblast
IC: intercalated cell
LOH: loop of Henle
MC: mesangial cell
MYL: myeloid cell
PC: principal cell
PEC: parietal epithelial cell
POD: podocyte
PT: proximal tubular epithelial cell
TAL: thick ascending loop of Henle
VSMC: vascular smooth muscle cell

