## Supplementary Materials for "Multi-scalar data integration links glomerular angiopoietin-tie signaling pathway activation with progression of diabetic kidney disease"

1. Supplementary acknowledgement – KPMP
2. Supplementary methods
3. Supplementary references
4. Supplementary tables:
    - Table S1. Proteins/SOMAmers associated with DKD progression
    - Table. S2 Association of plasma ANGPT2 measured by ELISA with risk of composite outcome
    - Table S3. ANG-TIE signaling pathway related genes from three database
    - Table S4. Basic clinical data of DKD patients included in the ScRNAseq analysis
5. Supplementary Figures: S1 to S6
    - Figure S1. Curation of an unbiased ANG-TIE signaling gene set.
    - Figure S2. Correlation matrix between the univariate significant plasma proteins and the lasso cross validation curve..
    - Figure S3. Time-dependent ROC curve truncated at 5 years for clinical model and the joint clinical and biomarker model.
    - Figure S4. Functional characterization of the ANG-TIE signaling network in the kidney.
    - Figure S5. Association of tubular ANG-TIE signaling pathway score with plasma ANGPT2 level and kidney outcome (n=25).
    - Figure S6. TEK gene expression in KPMP single cell data: DKD and living kidney donors (LD).

**Supplementary acknowledgement**

**<u>For the Kidney Precision Medicine Project</u>**

*American Association of Kidney Patients, Tampa, FL*: Richard Knight

*Beth Israel Deaconess, Boston, MA*: Stewart H. Lecker; Isaac Stillman

*Boston Cell Standards, Boston, MA*: Steve Bogen

*Boston University and Boston Medical Center, Boston, MA*: Afolarin A. Amodu; Titlayo Ilori; Shana Maikhor; Insa Schmidt, Laurence H. Beck; Joel M. Henderson; Ingrid Onul; Ashish Verma, Sushrut Waikar

*Brigham & Women's Hospital, Boston, MA*: Gearoid M. McMahon; M. Todd Valerius; Sushrut Waikar; Astrid Weins; Mia R. Colona

*Broad Institute, Cambridge, MA*: Anna Greka; Nir Hacohen; Paul J. Hoover; Jamie L. Marshall

*Case Western Reserve, Cleveland, OH*: Mark Aulisio; Yijiang M. Chen; Andrew Janowczyk; Catherine Jayapandian; Vidya S. Viswanathan; William S. Bush; Dana C. Crawford; Anant Madabhushi

*Cleveland Clinic, Cleveland, OH*: Lakeshia Bush; Leslie Cooperman; Agustin Gonzalez-Vicente; Leal Herlitz; Stacey Jolly; Jane Nguyen; John O'toole; Ellen Palmer; Emilio Poggio; John Sedor; Dianna Sendrey; Kassandra Spates-Harden; Jonathan Taliercio

*University of Colorado*, *Denver, CO*: Petter M. Bjornstad; Laura Pyle; Carissa Vinovskis

*Columbia University, New York, NY*: Paul Appelbaum; Jonathan M. Barasch; Andrew S. Bomback; Pietro A. Canetta; Vivette D. D'Agati; Krzysztof Kiryluk; Satoru Kudose; Karla Mehl; Ning Shang; Olivia Balderes

*Duke University, Durham, NC*: Shweta Bansal

*European Molecular Biology Laboratory, Heidelberg, Germany*: Theodore Alexandrov

*Harvard University, Cambridge, MA*: Helmut Rennke

*Indiana University, Indianapolis, IN*: Tarek M. El-Achkar; Daria Barwinska; Sharon Bledsoe; Katy Borner; Andreas Bueckle; Yinghua Cheng; Pierre C. Dagher; Kenneth W. Dunn; Michael T. Eadon; Michael J. Ferkowicz; Bruce W. Herr; Katherine J. Kelly; Ricardo Melo Ferreira; Ellen M. Quardokus; Elizabeth Record; Marcelino Rivera; Jing Su; Timothy A. Sutton; James C. Williams, Jr.; Seth Winfree

*John Hopkins University, Baltimore, MD*: Steven Menez; Chirag R. Parikh; Avi Rosenberg; Celia P. Corona-Villalobos; Yumeng Wen

*Joslin Diabetes Center, Boston, MA*: Camille Johansen; Sylvia E. Rosas; Neil Roy; Jennifer Sun;

Mark Williams

*Mount Sinai, New York, NY*: Evren U. Azeloglu; Jens Hansen; Cijang He; Ravi Iyengar; Yuguang Xiong

*Northshore, Evanston, IL*: Pottumarthi Prasad

*Northwestern University, Evanston, IL*: Anand Srivastava

*Ohio State University, Columbus, OH*: Sethu M. Madhavan; Samir Parikh; Brad Rovin; John P. Shapiro

*Pacific Northwest National Laboratories, Richland, WA*: Christopher R. Anderton; Jessica Lukowski; Ljiljana Pasa-Tolic; Dusan Velickovic

*Parkland Center for Clinical Innovation, Dallas, TX*: George (Holt) Oliver

*Patient Advocates*: Joseph Ardayfio; Jack Bebiak; Keith Brown; Taneisha Campbell; Catherine E. Campbell; Lynda Hayashi; Nichole Jefferson; Glenda V. Roberts; John Saul; Anna Shpigel; Christy Stutzke; Robert Koewler; Roy Pinkeney

*Princeton University, Princeton, NJ*: Rachel Sealfon; Olga Troyanskaya; Aaron Wong

*Providence Medical Research Center, Spokane, WA*: Katherine R. Tuttle

*Seattle Children's Hospital, Seattle, WA*: Ari Pollack

*Stanford University, Stanford, CA*: Yury Goltsev

*State University of New York, Buffalo, NY*: Nicholas Lucarelli; Pinaki Sarder

*University of California San Diego, La Jolla, CA*: Blue B. Lake; Kun Zhang

*University of California San Francisco, San Francisco, CA*: Patrick Boada; Zoltan G. Laszik; Garry Nolan; Kavya Anjani; Minnie Sarwal; Tariq Mukatash; Tara Sigdel

*University of Cincinnati, Cincinnati, OH*: Rita R. Alloway; Ashley R. Burg; Paul J. Lee; Adele Rike; Tiffany Shi; E. Steve Woodle

*University of Michigan, Ann Arbor, MI*; Ulysses GJ. Balis; Victoria M. Blanc; Ninive C. Conser; Sean Eddy; Renee Frey; Yougqun He; Jeffrey B. Hodgin; Matthias Kretzler; Chrysta Lienczewski; Jinghui Luo; Laura H. Mariani; Rajasree Menon; Edgar Otto; Jennifer Schaub; Becky Steck;

*University of Pittsburgh, Pittsburgh, PA*: Michele M. Elder; Matthew Gilliam; Daniel E. Hall; Raghavan Murugan; Paul M. Palevsky; Parmjeet Randhawa; Matthew Rosengart; Mitchell Tublin; Tina Vita; John A. Kellum; James Winters

*University of Washington, Seattle, WA*: Charles E. Alpers; Ashley Berglund; Kristina N. Blank; Jonas Carson; Stephen Daniel; Ian H. De Boer; Ashveena L. Dighe; Frederick Dowd; Stephanie M. Grewenow; Jonathan Himmelfarb; Andrew N. Hoofnagle; Christine Limonte; Robyn L. McClelland; Sean D. Mooney; Kasra Rezaei; Stuart Shankland; Jamie Snyder; Ruikang Wang;

Adam Wilcox; Kayleen Williams; Christopher Park

*UT Health San Antonio, San Antonio, TX*: Shweta Bansal; Richard Montellano; Annapurna Pamreddy; Kumar Sharma; Manjeri Venkatachalam; Hongping Ye; Guanshi Zhang

*UT Southwestern Medical Center, Dallas, TX*: S. Susan Hedayati; Asra Kermani; Simon C. Lee; Christopher Y. Lu; R. Tyler Miller; Orson W. Moe; Jiten Patel; Anil Pillai; Kamalanathan Sambandam; Jose Torrealba; Robert D. Toto; Miguel Vazquez; Nancy Wang; Natasha Wen; Dianbo Zhang; Harold Park

*Vanderbilt University, Nashville, TN*: Richard M. Caprioli; Nathan Patterson; Kavya Sharman; Jeffrey M. Spraggins; Raf Van de Plas

*Washington University in St. Louis, St. Louis, MO*: Jeanine Basta; Sabine M. Diettman; Joseph P. Gaut; Sanjay Jain; Michael I. Rauchman; Anitha Vijayan

*Yale University, New Haven, CT*: Lloyd G. Cantley; Vijaykumar R. Kakade; Dennis Moledina; Melissa M. Shaw; Ugochukwu Ugwuowo; Francis P. Wilson; Tanima Arora

### Supplementary Methods

#### *SOMAscan measurement*

Briefly, plasma samples were incubated with aptamers, washed and quantified with an Agilent microarray (Agilent Technologies). Normalization and calibration with replicate measurements of a common pooled calibrator plasma sample were carried out as previously described (S1). Samples in this study were analyzed in batches balanced by prospective case status and masked to the laboratory operators and data processing scientists. Twelve replicate samples across the batches allowed for universal calibration based on intercept and beta estimates from the linear regression model drawn from PROC GLM in SAS.

#### *Statistical analysis- Lasso Cox*

Lasso introduces a penalty on the regression coefficients, leading to some coefficients shrinking to zero and thereby simultaneously performing variable selection (S2). The penalty factor of the lasso Cox model was learned from a 5 fold 200 repeats cross validation process, based on minimizing the cross validated deviance defined as -2*log(partial likelihood) of the model. The repeated grid-search k-fold cross-validation procedure was selected due to the variability of lasso cross validation results in different data splits, which is exaggerated in small sample size and large candidate feature scenario. The method incorporated the variability of different data splits, thus improving the reliability and confidence of the selected optimal model (S3).

S3. Krstajic D, Buturovic LJ, Leahy DE, Thomas S: Cross-validation pitfalls when selecting and assessing regression and classification models. J Cheminform 2014;6

**Table S1. Proteins/SOMAmers associated with DKD progression**

| Protein | HR | P value | HR.confint.lower | HR.confint.upper |
|---|---|---|---|---|
| EGFR.2677.1.1 | 0.166 | 0.000 | 0.063 | 0.434 |
| APOA1.2750.3.2 | 0.266 | 0.001 | 0.118 | 0.598 |
| CTSV.3364.76.2 | 0.303 | 0.001 | 0.146 | 0.629 |
| CLEC4M.3030.3.2 | 0.168 | 0.002 | 0.055 | 0.510 |
| TNFRSF1A.2654.19.1 | 3.255 | 0.002 | 1.546 | 6.852 |
| CCL20.2468.62.3 | 2.173 | 0.004 | 1.283 | 3.678 |
| HAMP.3504.58.2 | 0.635 | 0.004 | 0.467 | 0.864 |
| IL22.2778.10.2 | 2.150 | 0.004 | 1.277 | 3.622 |
| IL1RL1.4234.8.2 | 2.744 | 0.004 | 1.375 | 5.476 |
| SERPINA4.3449.58.2 | 0.189 | 0.004 | 0.060 | 0.592 |
| PGLYRP1.3329.14.2 | 1.714 | 0.006 | 1.171 | 2.509 |
| CD5L.3293.2.4 | 2.158 | 0.006 | 1.252 | 3.718 |
| IL1RL2.2994.71.2 | 0.159 | 0.006 | 0.043 | 0.585 |
| ECE1.3611.70.4 | 0.399 | 0.007 | 0.205 | 0.773 |
| GSN.4775.34.3 | 0.146 | 0.007 | 0.036 | 0.586 |
| NTN4.3327.27.1 | 2.822 | 0.007 | 1.328 | 5.996 |
| FTH1.FTL.5934.1.3 | 0.643 | 0.008 | 0.465 | 0.890 |
| YWHAQ.7625.27.3 | 2.713 | 0.008 | 1.297 | 5.674 |
| DIABLO.3122.6.2 | 3.907 | 0.008 | 1.425 | 10.715 |
| GHR.2948.58.2 | 0.339 | 0.008 | 0.152 | 0.755 |
| IBSP.3415.61.2 | 1.862 | 0.008 | 1.174 | 2.951 |
| TNFRSF9.2598.9.3 | 2.077 | 0.009 | 1.204 | 3.581 |
| ANGPT2.2602.2.2 | 2.323 | 0.009 | 1.236 | 4.365 |
| TPM1.5033.27.1 | 1.902 | 0.009 | 1.171 | 3.091 |
| TNFRSF4.3730.81.2 | 6.035 | 0.009 | 1.553 | 23.446 |
| CA6.3352.80.3 | 0.503 | 0.010 | 0.298 | 0.847 |
| CSF3R.2719.3.4 | 1.679 | 0.010 | 1.133 | 2.490 |
| HTRA2.3317.33.1 | 2.322 | 0.011 | 1.217 | 4.432 |
| ANP32B.4194.26.3 | 3.512 | 0.011 | 1.337 | 9.222 |
| HSPB1.11103.24.3 | 0.450 | 0.011 | 0.243 | 0.834 |
| VTN.13125.45.3 | 0.451 | 0.011 | 0.244 | 0.836 |
| CKAP2.5345.51.3 | 0.394 | 0.012 | 0.191 | 0.811 |
| DDR1.4122.12.2 | 0.310 | 0.012 | 0.124 | 0.774 |
| CSF1.3738.54.1 | 2.516 | 0.013 | 1.217 | 5.203 |
| CD40LG.3534.14.2 | 1.521 | 0.018 | 1.074 | 2.153 |
| PPIB.4718.5.2 | 1.706 | 0.019 | 1.092 | 2.664 |
| NID2.3633.70.5 | 5.790 | 0.019 | 1.330 | 25.208 |
| AMN.4322.28.3 | 3.775 | 0.021 | 1.223 | 11.656 |
| REG1A.13095.51.3 | 2.046 | 0.021 | 1.114 | 3.755 |
| B2M.3485.28.2 | 3.331 | 0.022 | 1.191 | 9.317 |

| | | | | |
|---|---|---|---|---|
| NTRK1.3477.63.2 | 1.577 | 0.022 | 1.067 | 2.329 |
| ALB.3707.12.2 | 0.206 | 0.023 | 0.053 | 0.805 |
| THBS2.3339.33.1 | 1.614 | 0.025 | 1.062 | 2.452 |
| PDIA3.4719.58.2 | 0.205 | 0.025 | 0.051 | 0.822 |
| NOTCH1.5107.7.2 | 0.300 | 0.026 | 0.104 | 0.869 |
| TIMP1.2211.9.6 | 1.885 | 0.027 | 1.076 | 3.303 |
| CD27.5412.53.3 | 0.181 | 0.027 | 0.040 | 0.823 |
| ARID3A.3875.62.1 | 1.345 | 0.027 | 1.034 | 1.749 |
| SCARF1.5129.12.3 | 4.621 | 0.030 | 1.158 | 18.438 |
| CCL19.4922.13.1 | 1.726 | 0.031 | 1.052 | 2.832 |
| FGF8.4394.71.2 | 1.494 | 0.033 | 1.034 | 2.158 |
| IL1B.3037.62.1 | 1.607 | 0.033 | 1.039 | 2.486 |
| AIMP1.2714.78.2 | 0.315 | 0.033 | 0.109 | 0.914 |
| SHC1.5272.55.2 | 2.222 | 0.034 | 1.063 | 4.646 |
| HIPK3.3443.61.2 | 0.414 | 0.035 | 0.182 | 0.938 |
| CCL2.2578.67.2 | 1.291 | 0.035 | 1.018 | 1.636 |
| CD300C.5066.134.3 | 2.578 | 0.035 | 1.068 | 6.222 |
| NGF.5801.72.3 | 1.350 | 0.035 | 1.021 | 1.785 |
| IFNL1.4396.54.1 | 1.432 | 0.036 | 1.024 | 2.001 |
| ACP5.3232.28.2 | 0.297 | 0.036 | 0.096 | 0.924 |
| LCMT1.4237.70.3 | 0.139 | 0.036 | 0.022 | 0.882 |
| MAPK12.5005.4.1 | 1.972 | 0.036 | 1.044 | 3.727 |
| KDR.3651.50.5 | 0.247 | 0.038 | 0.066 | 0.926 |
| C7.2888.49.2 | 3.563 | 0.039 | 1.065 | 11.917 |
| FGF2.3025.50.1 | 0.682 | 0.040 | 0.474 | 0.982 |
| TNFRSF1B.3152.57.1 | 2.506 | 0.040 | 1.042 | 6.028 |
| NCAM1.4498.62.2 | 0.326 | 0.041 | 0.112 | 0.953 |
| CTSF.9212.22.3 | 0.326 | 0.041 | 0.112 | 0.954 |
| ESAM.2981.9.3 | 4.009 | 0.041 | 1.056 | 15.225 |
| IFNGR2.9180.6.3 | 2.181 | 0.041 | 1.031 | 4.616 |
| RPS3A.5484.63.3 | 0.436 | 0.042 | 0.196 | 0.970 |
| APCS.2474.54.5 | 0.410 | 0.043 | 0.173 | 0.971 |
| S100A4.14116.129.3 | 0.424 | 0.043 | 0.184 | 0.975 |
| TNFAIP6.5036.50.1 | 3.088 | 0.044 | 1.029 | 9.264 |
| ALCAM.5451.1.3 | 0.381 | 0.046 | 0.147 | 0.985 |
| GFRA3.2505.49.3 | 1.474 | 0.047 | 1.005 | 2.160 |
| FCGR2B.3310.62.1 | 1.651 | 0.048 | 1.005 | 2.710 |
| KLRK1.3056.11.1 | 2.939 | 0.048 | 1.011 | 8.546 |
| PIGR.3216.2.2 | 1.561 | 0.048 | 1.004 | 2.426 |
| CLEC11A.4500.50.2 | 2.554 | 0.048 | 1.008 | 6.474 |
| EHMT2.5843.60.3 | 0.391 | 0.049 | 0.154 | 0.995 |
| KLK6.3450.4.2 | 0.404 | 0.049 | 0.164 | 0.996 |
| SNRPF.5494.52.3 | 0.259 | 0.050 | 0.067 | 0.997 |
| PGAM1.3896.5.2 | 1.224 | 0.050 | 1.000 | 1.498 |

| Table. S2 Association of plasma ANGPT2 measured by ELISA with risk of composite outcome | | | |
|---|---|---|---|
| **ANGPT2(pg/ml)** | **Event (n/N)** | **Hazard ratio (95% Cl)** | **P-value** |
| log2ANGPT2 | 28/58 | 2.11 (1.30-3.41) | 0.002 |
| Q1(1005-1767) | 3/15 | 1.00 (Ref) | |
| Q2(1813-2333) | 7/14 | 5.74 (1.38-23.89) | 0.016 |
| Q3(2345-3393) | 7/14 | 4.99 (1.27-19.55) | 0.021 |
| Q4(3416-8662) | 11/15 | 8.04 (2.13-30.38) | 0.002 |

| Table. S3 ANG-TIE signaling pathway related genes from three databases | | |
|---|---|---|
| PID_ANGIOPOIETIN_RECEPTOR_PATHWAY | REACTOME_TIE2_SIGNALING | NETPATH_TEK_SIGNALING |
| AGTR1 | ANGPT1 | ACTB |
| AKT1 | ANGPT2 | BIRC5 |
| ANGPT1 | ANGPT4 | EEFSEC |
| ANGPT2 | DOK2 | GRB7 |
| ANGPT4 | GRB14 | ITGB1 |
| BMX | GRB2 | MASP2 |
| CDKN1A | GRB7 | PIK3R2 |
| CRK | HRAS | PXN |
| DOK2 | KRAS | SPHK1 |
| ELF1 | NRAS | WAS |
| ELF2 | PIK3CA | ADAM30 |
| ELK1 | PIK3CB | BMX |
| ETS1 | PIK3R1 | EGFR |
| F2 | PIK3R2 | GSK3B |
| FES | PTPN11 | ITGB2 |
| FGF2 | SHC1 | MMP2 |
| FN1 | SOS1 | PITPNM3 |
| FOXO1 | TEK | RAC1 |
| FYN | | SRC |
| GRB14 | | WASF1 |
| GRB2 | | ADSS |
| GRB7 | | CAPZB |
| ITGA5 | | EIF3F |
| ITGB1 | | GTPBP3 |
| MAPK1 | | ITGB3 |
| MAPK14 | | MTOR |
| MAPK3 | | PKD1 |
| MAPK8 | | RAF1 |
| MMP2 | | STAT1 |
| NCK1 | | ZAP70 |
| NFKB1 | | AKT1 |
| NOS3 | | CBL |
| PAK1 | | EIF3I |
| PIK3CA | | GYS1 |
| PIK3R1 | | ITGB5 |
| PLD2 | | NCK1 |
| PLG | | PKD2 |
| PTK2 | | RASA1 |
| PTPN11 | | STAT3 |
| PXN | | ZNF397 |
| RAC1 | | AKT2 |

| | | |
|---|---|---|
| RASA1 | | CCT5 |
| RELA | | ELK1 |
| RPS6KB1 | | HAGH |
| SHC1 | | LIMCH1 |
| STAT5A | | NF1 |
| STAT5B | | PLA2G5 |
| TEK | | RELA |
| TNF | | STAT5A |
| | | ALB |
| | | CDC42 |
| | | F13A1 |
| | | HDAC7 |
| | | LONP1 |
| | | NFKBIA |
| | | PLCG1 |
| | | RHOA |
| | | STAT5B |
| | | ALDOA |
| | | CDH5 |
| | | FES |
| | | HIF1A |
| | | MAP2K1 |
| | | NONO |
| | | PLD1 |
| | | ROCK1 |
| | | STAT6 |
| | | ANGPT1 |
| | | CREB1 |
| | | FOXO1 |
| | | HNRNPF |
| | | MAP2K2 |
| | | NOS3 |
| | | PLD2 |
| | | RPS6KA1 |
| | | TEK |
| | | ANGPT2 |
| | | CRK |
| | | FOXO3 |
| | | HNRNPH1 |
| | | MAP2K4 |
| | | PAK1 |
| | | PLG |
| | | RPS6KA5 |
| | | TIE1 |
| | | ANGPT4 |
| | | CTNNB1 |
| | | FYN |

| | | |
|---|---|---|
| | | HNRNPM |
| | | MAP3K3 |
| | | PDCD6IP |
| | | PPP2CA |
| | | RPS6KB1 |
| | | TLN1 |
| | | ANGPTL1 |
| | | CTPS1 |
| | | GAPDH |
| | | HSP90AA1 |
| | | MAPK1 |
| | | PDIA3 |
| | | PRDX4 |
| | | RUVBL2 |
| | | TNIP2 |
| | | ANKRD28 |
| | | DIAPH1 |
| | | GCDH |
| | | HSPD1 |
| | | MAPK14 |
| | | PDPK1 |
| | | PRKCZ |
| | | SELP |
| | | TXNRD1 |
| | | ARHGAP5 |
| | | DOK2 |
| | | GNA11 |
| | | ILK |
| | | MAPK3 |
| | | PECAM1 |
| | | PTK2 |
| | | SERPINH1 |
| | | TXNRD3 |
| | | BCAR1 |
| | | DOK4 |
| | | GRB14 |
| | | ITGA5 |
| | | MAPK8 |
| | | PGK1 |
| | | PTPN11 |
| | | SHC1 |
| | | VEGFA |
| | | BCL2 |
| | | DYNC1H1 |
| | | GRB2 |
| | | ITGAV |
| | | MAPK9 |

| | | |
|---|---|---|
| | | PIK3R1 |
| | | PTPRB |
| | | SOS1 |
| | | VIM |

| Table. S4 Basic clinical data of DKD patients included in the ScRNAseq analysis | | | | | | | |
|---|---|---|---|---|---|---|---|
| **Tissue Type** | **Sex** | **Age(Years)** | **Race** | **Baseline eGFR (ml/min/1.73m2)** | **Diabetes** | **Diabetes Duration (Years)** | **Hypertension History** |
| CKD | Female | 60-69 | White,Other | 20-29 | Yes | 20-24 | Yes |
| CKD | Female | 60-69 | Black or African-American | 80-89 | Yes | 5-9 | Yes |
| CKD | Female | 30-39 | White | 100-109 | Yes | 20-24 | Yes |
| CKD | Female | 60-69 | Black or African-American | 110-119 | Yes | 5-9 | Yes |
| CKD | Male | 70-79 | | 40-49 | Yes | 10-14 | Yes |
| CKD | Female | 30-39 | White | 30-39 | Yes | 20-24 | Yes |
| CKD | Male | 60-69 | White | 30-39 | Yes | 0-4 | No |
| CKD | Female | 70-79 | White | 60-69 | Yes | 10-14 | Yes |
| CKD | Male | 70-79 | White | 40-49 | Yes | 25-29 | Yes |
| CKD | Female | 60-69 | White | 40-49 | Yes | 30-34 | Yes |

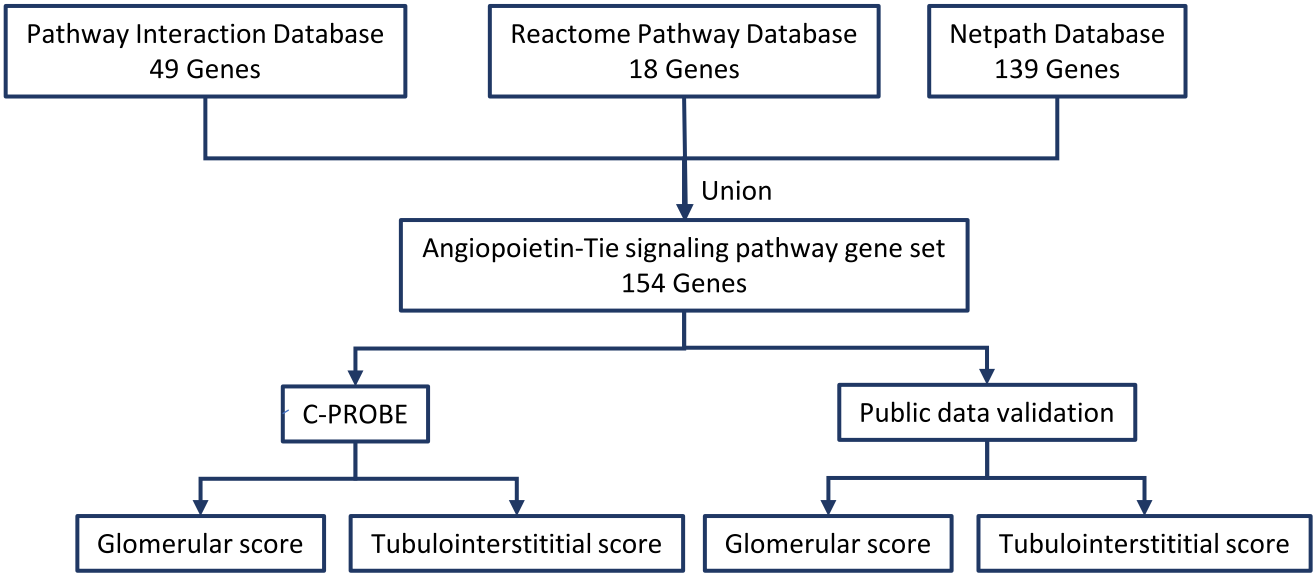


**Figure S1. Curation of an unbiased ANG-TIE signaling gene set.**

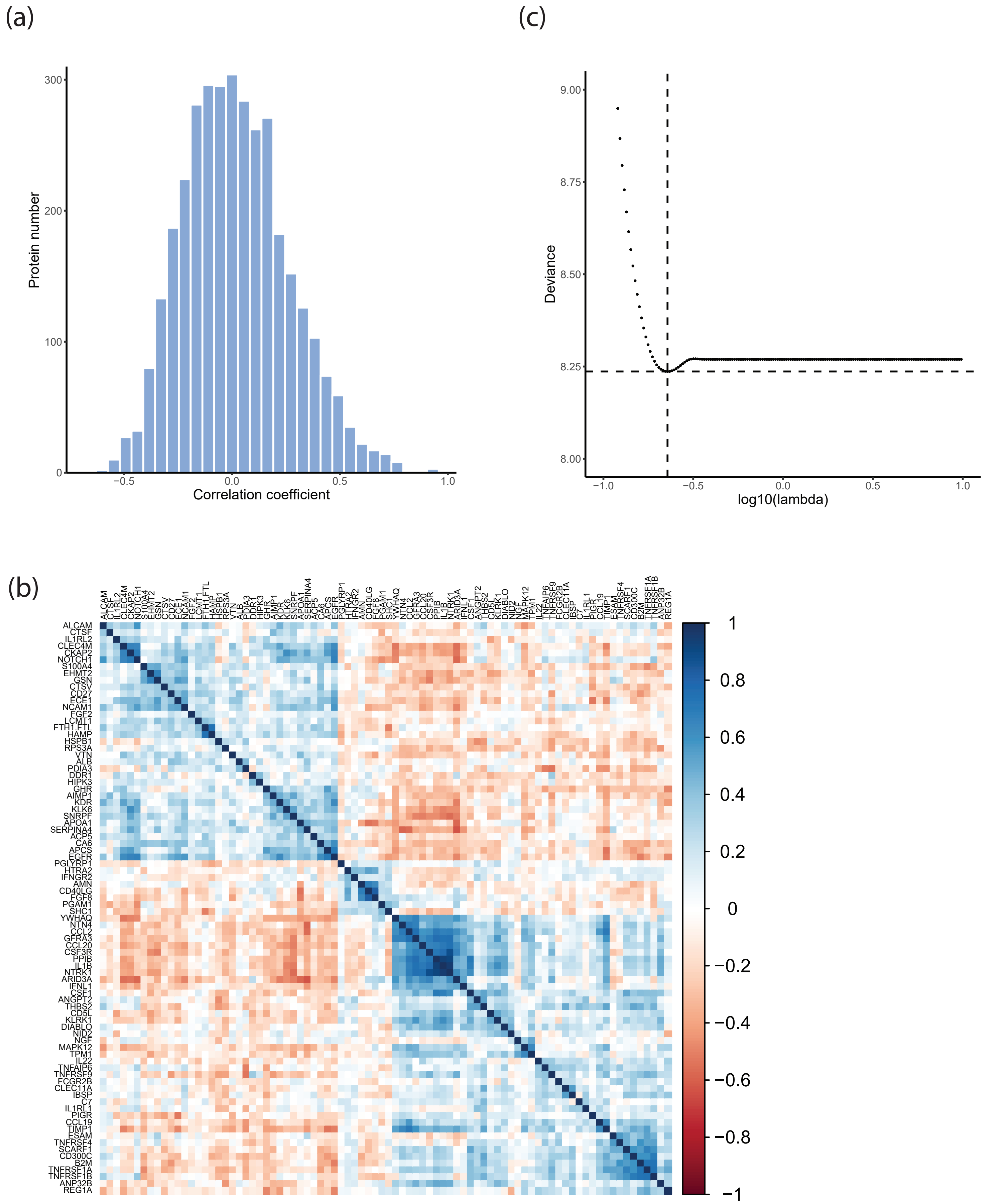


**Figure S2. Correlation matrix between the univariate significant plasma proteins and the lasso cross validation curve.** (a) Correlation distribution of the univariate significant plasma proteins. (b) Correlation structure of univariate significant proteins. Proteins were ordered by hierarchical clustering. Distinct correlation patterns were observed for plasma proteins in different clusters. (c) Cross validation curve of the lasso Cox model. The cross validation curve was the average of 200 repeats. A set of 3 biomarkers were selected based on the tuning parameter log(lambda)= -0.64 that gave the lowest deviance.

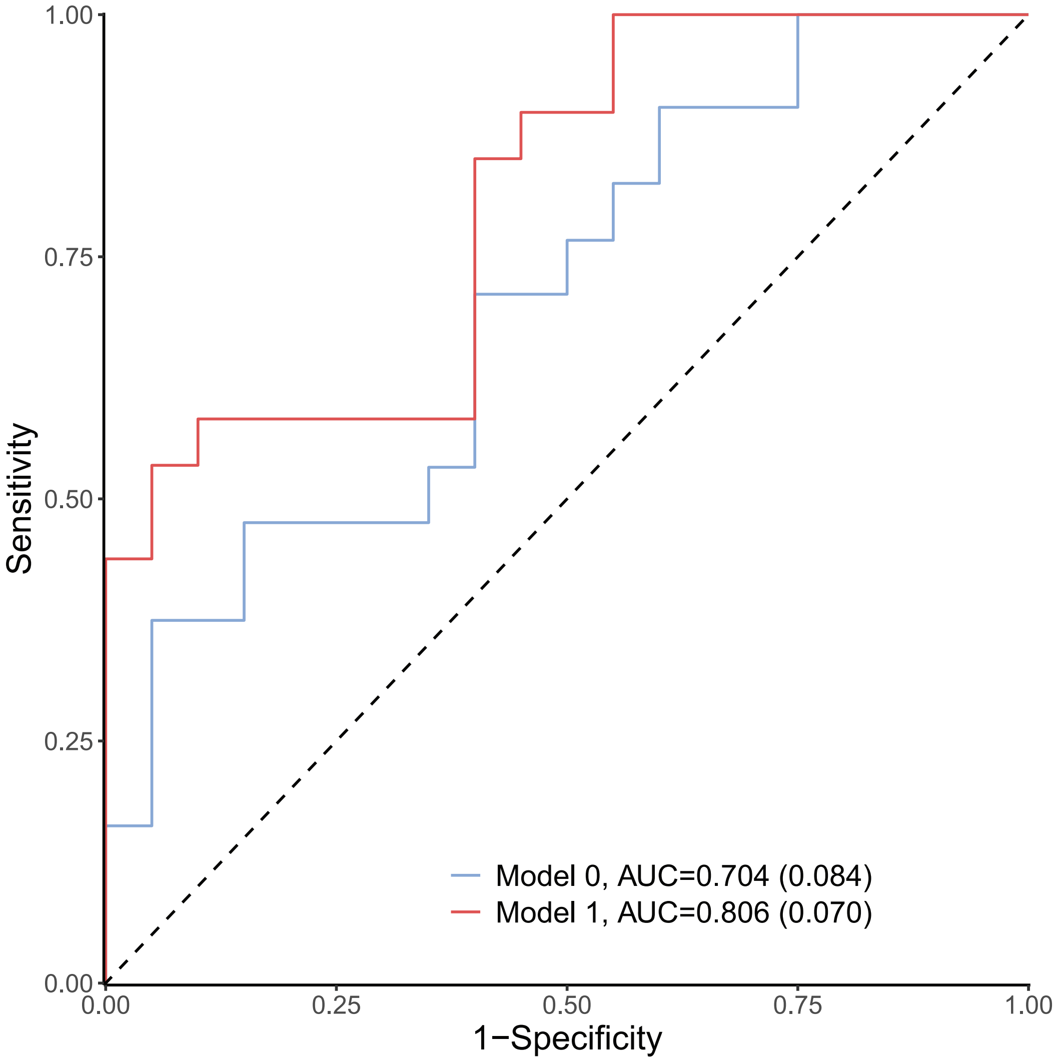


**Figure S3. Time-dependent ROC curve truncated at 5 years for clinical model and the joint clinical and biomarker model.**

Model 0 covariates: age, gender, race, eGFR and uACR.

Model 1 covariates: age, gender, race, eGFR uACR, ANGPT2, CLEC4M and EGFR.

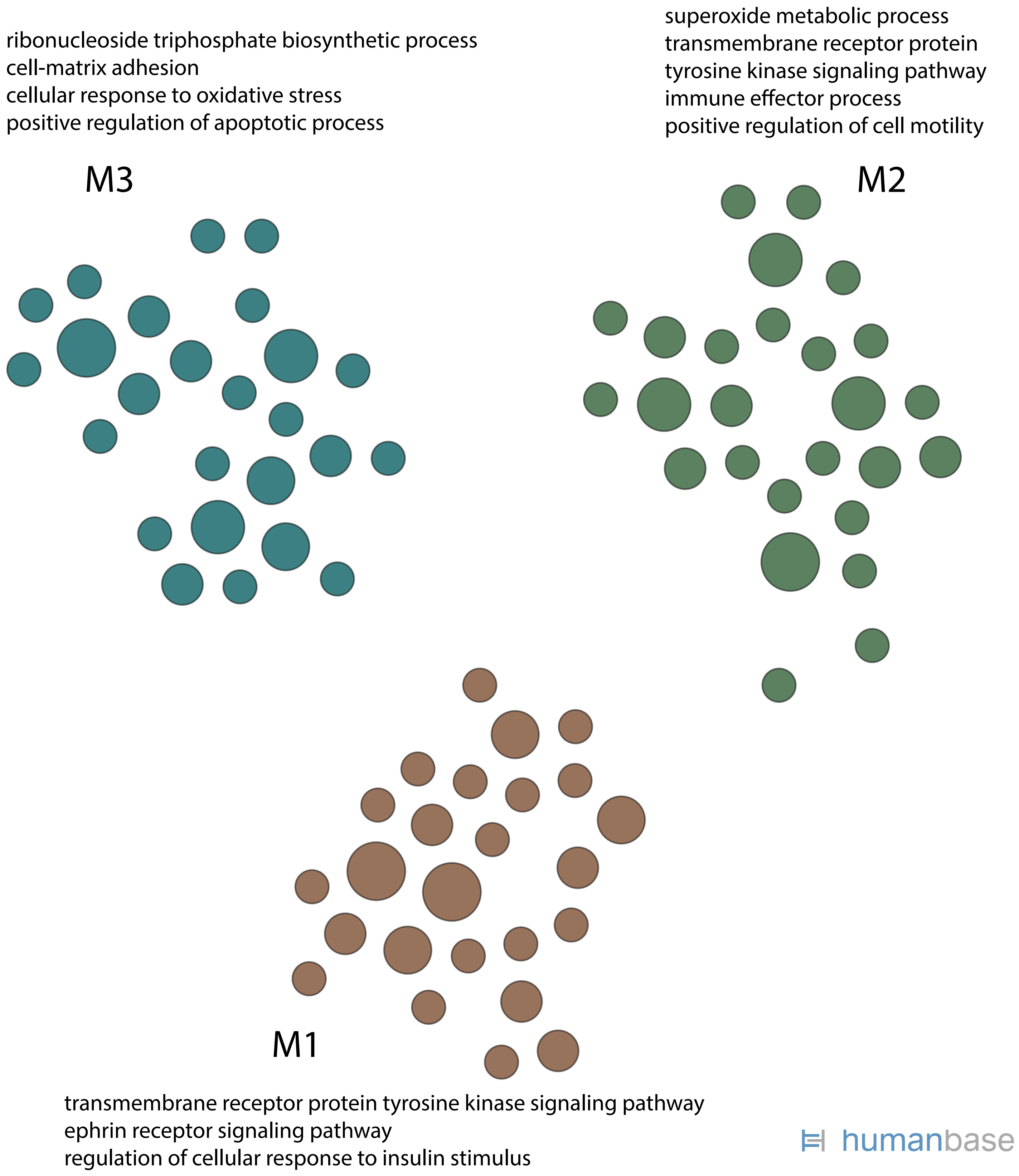


**Figure S4. Functional characterization of the curated ANG-TIE signaling network genes in the kidney.** Kidney functional modules were generated using HumanBase (https://hb.flatironinstitute.org/), by projecting the ANG-TIE signature onto the kidney active functional network followed by community clustering to identify cohesive functional modules. Enriched biological processes are shown for each module.

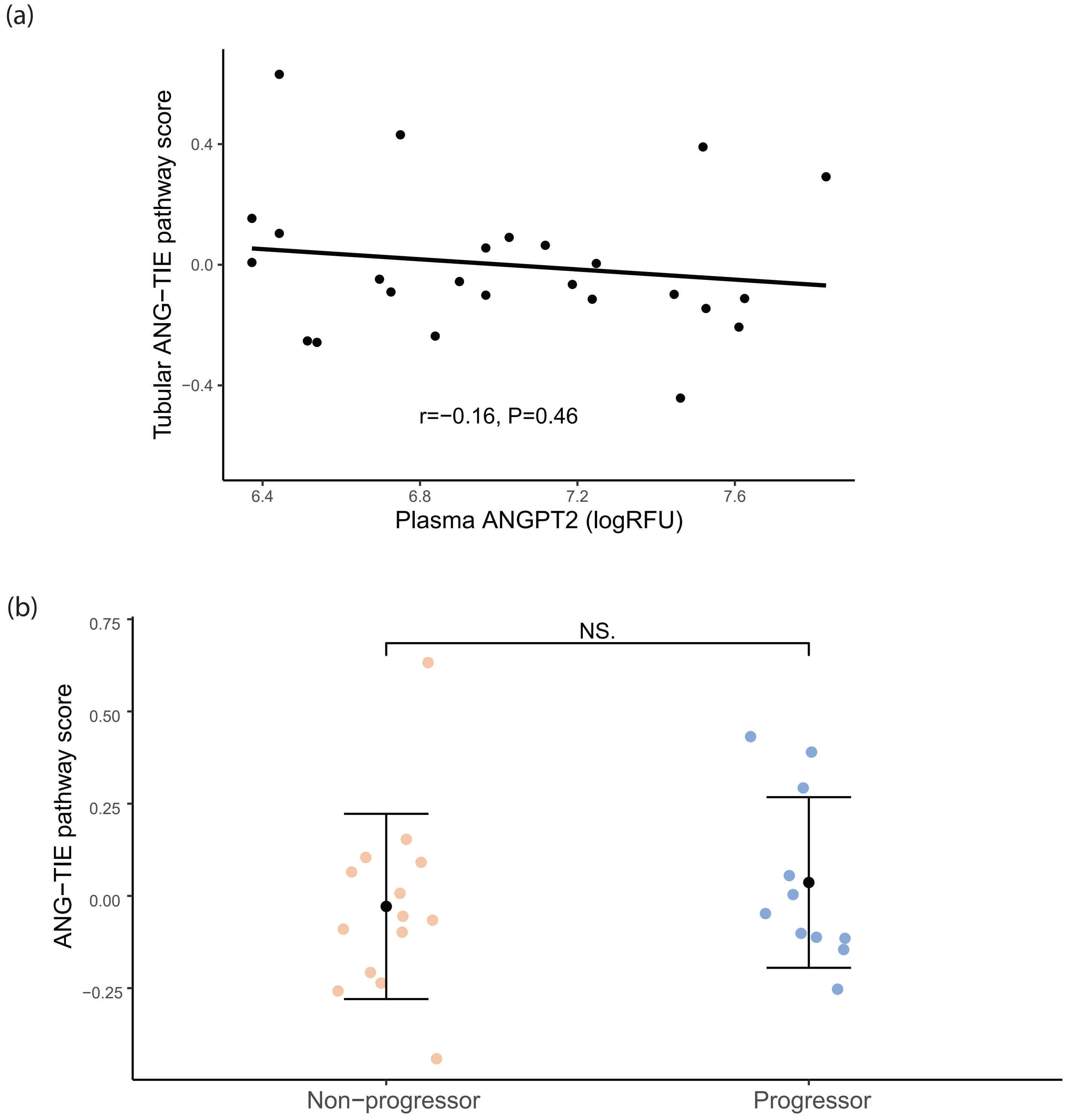


**Figure S5. Association of tubular ANG-TIE pathway activation score with plasma ANGPT2 level and kidney outcome (n=25).** (a) The association of plasma ANGPT2 level with tubular ANG-TIE pathway activation score (n=25). (b) Tubular ANG-TIE pathway activation score association with outcome in patients from C-PROBE Group B. (NS indicates p value > 0.05).

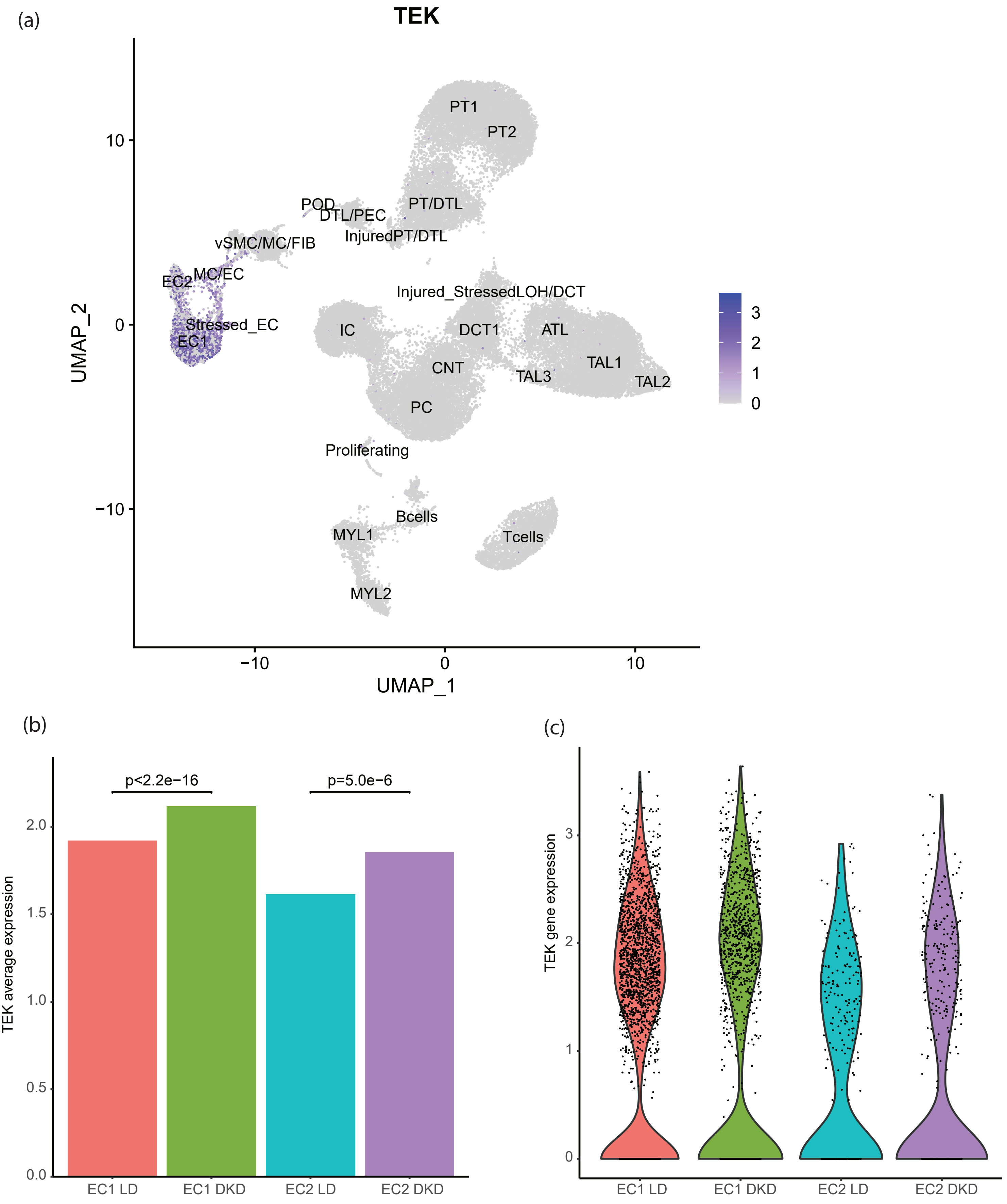


**Figure S6. *TEK* gene expression in KPMP single cell data: DKD and living kidney donors (LD).** (a) UMAP of *TEK* gene expression in KPMP data. Cells clusters labeled by cell identity. (b) Average value of *TEK* gene in endothelial cells whose *TEK* expression is more than 0. Mann–Whitney U test was used to test the difference between groups. (c) Violin plot of *TEK* gene in endothelial cells
